# Colchicine for Prevention of Major Adverse Cardiovascular Events: A systematic review and meta-analysis of randomized clinical trials

**DOI:** 10.1101/2024.12.19.24319310

**Authors:** Federico Ballacci, Federica Giordano, Cristina Conte, Alessandro Telesca, Valentino Collini, Massimo Imazio

## Abstract

**Aims:** Inflammation is a main pathophysiological driver in atherosclerotic cardiovascular diseases (ASCVD). Low-dose long-term colchicine for secondary prevention in patients with established ASCVD has been studied in multiple randomized trials in the last decade.

This meta-analysis aimed to evaluate the efficacy and safety of long-term low-dose colchicine for secondary prevention in patients with established ASCVD.

**Methods:** We conducted a systematic review and meta-analysis following PRISMA guidelines to evaluate studies reporting long-term outcomes in patients with ASCVD. We systematically searched PubMed, EMBASE and Scopus databases for relevant studies up to December 1, 2024. The primary outcome was the occurrence of Major Adverse Cardiovascular Events (MACE), a composite of cardiovascular death (CVD), myocardial infarction (MI) and stroke. Random-effects models were used to calculate pooled risk ratios (RR).

**Results:** Ten randomized clinical trials enrolling 22532 patients were identified. Addition of colchicine to standard medical treatment in patients with established ASCVD reduced the risk for MACE by 27% (RR 0.73, 95% CI 0.57 – 0.95), with a number needed to treat of 52. Colchicine was found to significantly reduce the risk of myocardial infarction (RR 0.83, 95% CI 0.72 - 0.96) and coronary revascularization (RR 0.79, 95% CI 0.65 - 0.94). There were no significant differences between the two groups concerning cardiovascular and non-cardiovascular mortality, risk of serious gastrointestinal events, infections requiring hospitalization and cancer.

**Conclusions:** These findings support the use of long-term low-dose colchicine for secondary prevention of MACE in clinical practice.

## Introduction

Atherosclerotic cardiovascular disease (ASCVD) is the leading cause of mortality worldwide, with a rising global burden despite advances in prevention and treatment strategies.

Current guidelines strongly emphasize secondary prevention measures after coronary artery disease or stroke diagnosis, including lifestyle interventions, risk factor control, and antithrombotic therapy.[1,2] However, a significant residual risk of major adverse cardiovascular events (MACE) persists even with optimal treatment adherence.[3,4]

Inflammation is a key pathophysiological driver of atherosclerosis, with the potential to be an actionable target, as suggested by the association between inflammatory biomarkers, anti-inflammatory therapy and cardiovascular outcomes.[5–8] Colchicine, a potent and widely available anti-inflammatory agent extracted from plants of the genus Colchicum, has been widely used in the treatment of inflammatory diseases such as gout, familial Mediterranean fever, and pericarditis with good efficacy and safety profiles.[9–15] Its anti-inflammatory effects involve multiple pathways, including inhibition of microtubule formation and neutrophil function (i.e. chemotaxis, adhesion and mobilization), as well as inhibition of NLRP3 inflammasome.[16]

Early landmark studies, such as the COLCOT and the LoDoCo2 trials, demonstrated that colchicine reduces cardiovascular events in coronary artery disease.[17–20] However, these initial results have been challenged by subsequent trials, including CONVINCE in stroke patients[21] and CLEAR in post-myocardial infarction patients,[22] which have shown neutral results. To address these uncertainties, we conducted a systematic review and meta-analysis of randomized clinical trials to assess the overall effect of colchicine on major adverse cardiovascular events and their individual components in patients with coronary disease, recent myocardial infarction, and stroke.

## Methods

This study was performed in accordance with the Preferred Reporting Items for Systematic Reviews and Meta-analysis (PRISMA) Guidelines.[23] The study protocol and the analysis plan were developed prospectively, and are available on the Open Science Framework (https://osf.io/dqgwj).

### Search strategy

A comprehensive search for relevant articles on PubMed, EMBASE and Scopus was performed from inception through December 1^st^, 2024.

The search strategy combined the term “colchicine” with cardiovascular-related terms including “atherosclerosis”, “coronary artery disease”, “coronary heart disease”, “angina”, “infarction”, “coronary syndrome”, “ACS”, “ischemic heart disease”, “percutaneous coronary intervention”, “PCI”, “myocardial ischemia” and “stroke”, with appropriate adaptations to each database. Only full-text publications were considered eligible, while conference abstracts were excluded. No other exclusion criterion was used. Reference lists of included articles were screened to identify further relevant studies. For trials with multiple published reports, the report with the longest pre-specified follow-up was selected, provided that blinding integrity was maintained.

### Selection criteria

Studies were considered eligible if they were randomized clinical trials enrolling adult participants with established ASCVD, with the intervention consisting of long-term colchicine treatment (>3 months) compared to standard treatment. Studies were not considered eligible if they did not report outcomes for cardiovascular death or myocardial infarction. Reviews and case reports were excluded. Complete inclusion and exclusion criteria can be found in the **Supplemental Material**.

### Data extraction and quality assessment

Two investigators (FB and FG) independently reviewed the titles, abstracts, and full texts for inclusion criteria. Conflicts between reviewers were resolved by consensus or by consultation with a third investigator (MI). The following data were extracted from the included articles: first author or trial name, publication year, patient characteristics, intervention and follow-up duration. Risk of bias was independently assessed by two investigators (FB and FG) according to the Cochrane Collaboration revised risk-of-bias tool for randomized trial (Rob2 version 2.0) composed of five domains: randomization process (D1); deviations from intended interventions (D2); missing out-come data (D3); measurement of the outcome (D4); and selection of the reported result (D5).[24,25]

### Outcome definition

The primary endpoint was defined as the composite of myocardial infarction, stroke, and cardiovascular death (MACE). Secondary endpoints included the composite of myocardial infarction, stroke, cardiovascular death, and coronary revascularization, as well as each of these components analysed individually.

Safety outcomes that were consistently reported across the included studies were also evaluated (non-cardiovascular death, serious infections, pneumonia requiring hospitalization, serious adverse gastrointestinal events, cancer).

### Statistical Analysis

We used a random-effects model with the Paule-Mandel estimator to calculate the pooled risk ratios (RR) with 95% confidence intervals (CIs) for the primary and secondary outcomes. A fixed-effects model was also conducted as sensitivity analysis. Heterogeneity was assessed using the I² statistic, Galbraith plots were evaluated to identify outliers when I² exceeded 50%.

A pre-specified subgroup analysis was performed for studies exclusively enrolling patients with acute coronary syndromes (ACS). Meta-regression analyses were restricted to studies with complete covariate data, and were used to explore the relationship between treatment effects and study-level characteristics including the proportion of ACS patients, diabetes, female sex, statin use, active smoking, and beta-blocker therapy. Leave-one-out sensitivity analyses were performed to assess the robustness of the findings. Publication bias was evaluated by examination of funnel plots. All statistical analyses followed the intention-to-treat principle. Statistical significance was set at p < 0.05 (two-tailed). All analyses were performed using R version 4.3.3 (R Foundation for Statistical Computing, Vienna, Austria) with the *meta* and *metafor* packages.

## Results

### Search Results and Baseline Characteristics

The literature search identified a total of 5982 titles. After duplicate removal, titles and abstracts of 3284 papers were evaluated, and 43 articles were considered for full-text screening. Of these, 10 studies met the inclusion criteria and were included in the meta-analysis.[17,18,21,22,26–31] The references of all the included studies were searched for eligible entries, and no other study was deemed eligible for inclusion. The PRISMA flow diagram and checklist are shown in **Supplementary Table S1 and Figure S1**.

Seven studies were rated as being at “low” risk of bias, 1 study was rated as being at “high” risk of bias,[27] while 2 studies had unclear risk of bias[22,31] (**Supplementary Figure S2).**

A total of 22532 patients (female 4574, 20.3%) from 10 randomized clinical trials were included in the analysis. Trial inclusion criteria and comprehensive data are reported in **Table 1** and **Table 2**. In three trials, patients in the control group received optimal medical therapy without administration of placebo.[18,21,27] Differently from the other trials which included patients affected by coronary syndromes, one trial included patients affected by stroke as the main inclusion criterion.[21] Among coronary syndrome trials, 13,154 patients had ACS as a qualifying event, while 6,189 patients were affected by stable coronary artery disease.

**Table 1:** Key features of randomized trials included in this meta-analysis.

| Study | Year | Inclusion criteria | Intervention | Placebo-<br>controlle<br>d | Number of patients |  | Months<br>of<br>follow-up<br>(median) |
| --- | --- | --- | --- | --- | --- | --- | --- |
|  |  |  |  |  | Interventio<br>n group | Control<br>group |  |
| CONVINCE[21] | 2024 | Patients aged at least 40 years, with non-severe ischaemic stroke or high-risk transient ischaemic attack | Colchicine 0.5 mg daily | No | 1569 | 1575 | 34 |
| CLEAR[22] | 2024 | Patients aged $\geq 18$ with STEMI and those with NSTEMI plus risk factors | Colchicine 0.5 mg daily <sup>a</sup> | Yes | 3528 | 3534 | 36 |
| COCOMO-ACS[26] | 2024 | Age 18-82 years, with a coronary angiogram within 72 hours of NSTEMI diagnosis | Colchicine 0.5 mg daily | Yes | 30 | 31 | 18 |
| Radwan et al.[27] | 2024 | STEMI patients with successful primary PCI | Colchicine 0.5 mg daily | No | 100 | 100 | 6 |
| COPS[29] | 2021 | Patients aged 18-85 with ACS | Colchicine 0.5 mg twice daily for 30 days,<sup>b</sup> followed by 0.5 mg once daily for 11 months | Yes | 396 | 399 | 24 [32]‡ |
| Akrami et al.[28] | 2021 | Patients aged 40-70 years, with ACS | Colchicine 0.5 mg daily | Yes | 120 | 129 | 6 |
| LoDoCo2[18] | 2020 | Age 35-85 years, with proven coronary artery disease (proven by either angiography or non-invasive imaging, or CAC ≥ 400) with minimum 6-month clinical stability | Colchicine 0.5 mg daily after a 30-day run-in phase | Yes | 2762 | 2760 | 29 |
| COLCOT[17] | 2019 | Patients aged ≥ 18, with MI in the 30 | Colchicine 0.5 mg daily | Yes | 2366 | 2379 | 22.6 |
|  |  | days prior to enrolment, with completion of planned percutaneous revascularization procedures |  |  |  |  |  |
| LoDoCo[30] | 2013 | Age 35-85 years, angiographically proven coronary disease with minimum 6-month clinical stability | Colchicine 0.5 mg daily | No | 282 | 250 | 36 |
| Deftereos et al.[31] | 2013 | Diabetic patients aged 40-80 years, undergoing PCI | Colchicine 0.5 mg twice daily | Yes | 112 | 110 | 6 |

<sup>a</sup> Dose changed after interim analysis showed higher-then-expected discontinuation; <sup>b</sup> Dose reduction
allowed according to patients' tolerance.
‡ = MACE outcome data based on pre-specified extended follow-up.
ACS = acute coronary syndrome; PCI = percutaneous coronary intervention.

**Table 2.**
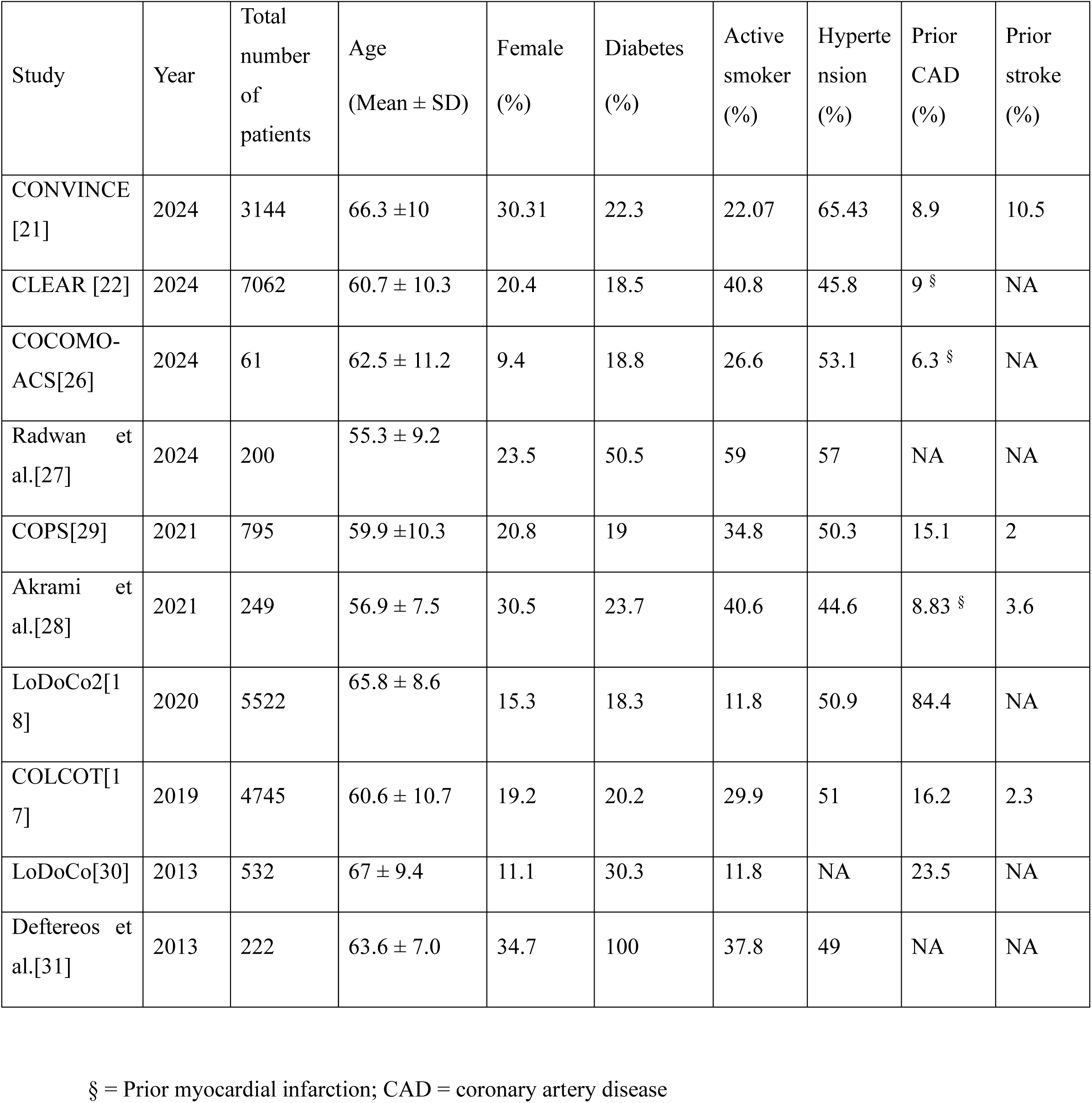
Demographics.

Median duration of follow-up ranged from a minimum of 6 months to a maximum of 36 months. In 6 trials, colchicine was administered at a dosage of 0.5 mg once daily,[17,21,26–28,30] while it was administered twice daily by Deftereos et al.[31] To ascertain treatment tolerance, investigators of the LoDoCo2 trial employed an open-label 30-day run-in phase before blinding and proceeding to the administration of 0.5 mg daily.[18] Patients enrolled in COPS the trial would receive colchicine 1 mg daily for the first month, followed by 0.5 mg once daily for 11 months.[29] The protocol of the CLEAR trial was changed due to higher-than-expected discontinuation rates, and the administered dosage was changed to 0.5 mg per day. [22]

Results of the COPS trial for the composite of MACE and coronary revascularization, as well as cardiovascular and non-cardiovascular death, are reported according to the two-year follow-up prespecified analysis. [29,32]

### Composite of Cardiovascular Death, Myocardial Infarction and Stroke

Seven studies reported data on the primary outcome of this meta-analysis.[17,18,21,22,26,28,30] A total of 1428 (6.5%) patients developed MACE. The pooled analysis showed that colchicine reduced the RR for MACE by 27% [95% confidence interval (CI) 0.57 – 0.95, I² = 58%] (**Figure 1**, **Figure 2**). Results were consistent for all studies at leave-one-out analysis. The estimated number needed to treat to prevent one occurrence of MACE was 52.

**Figure 1.**
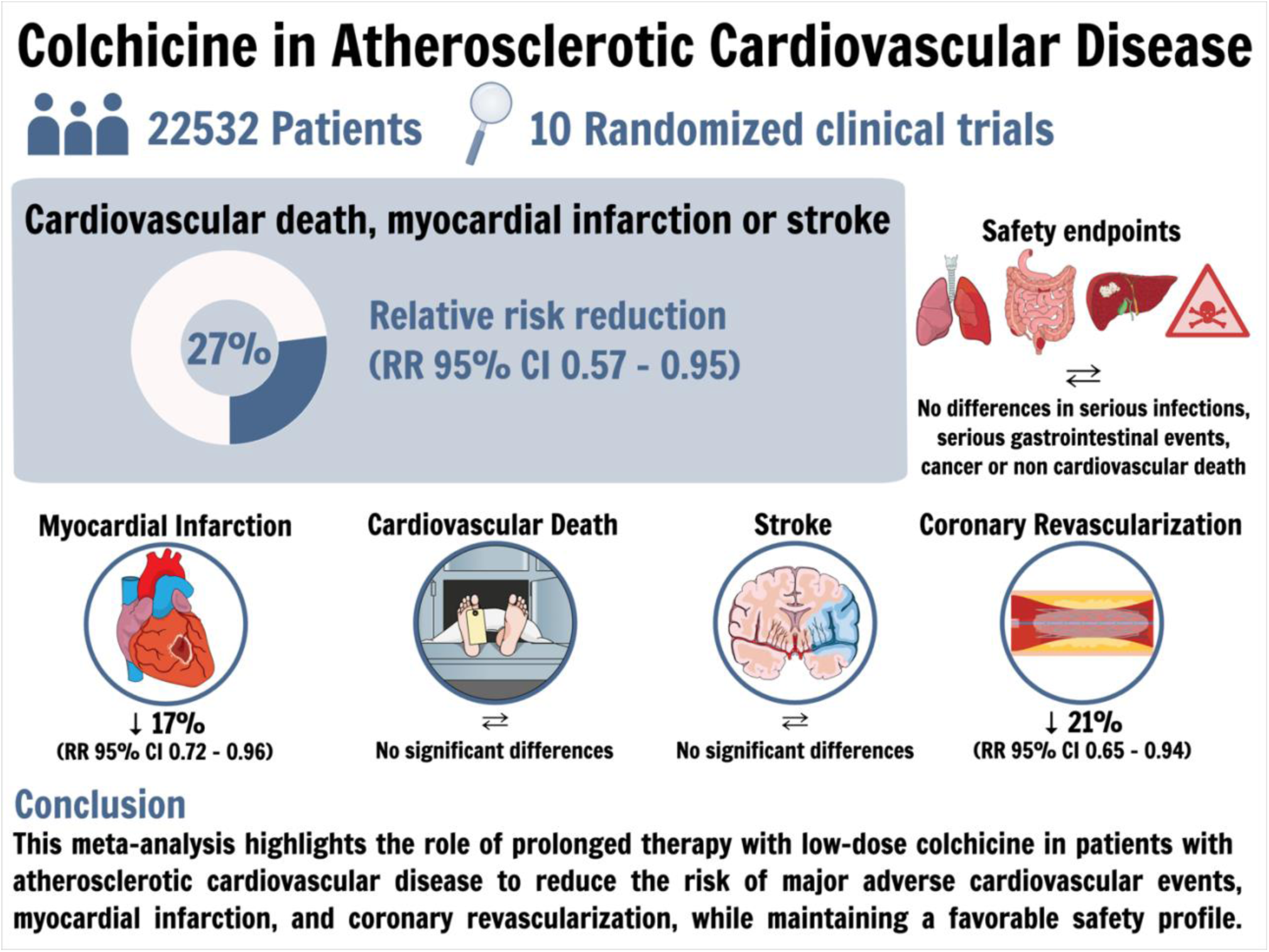
Graphical abstract: Main findings of the meta-analysis. RR = Risk Ratio; CI = Confidence Interval.

**Figure 2.**
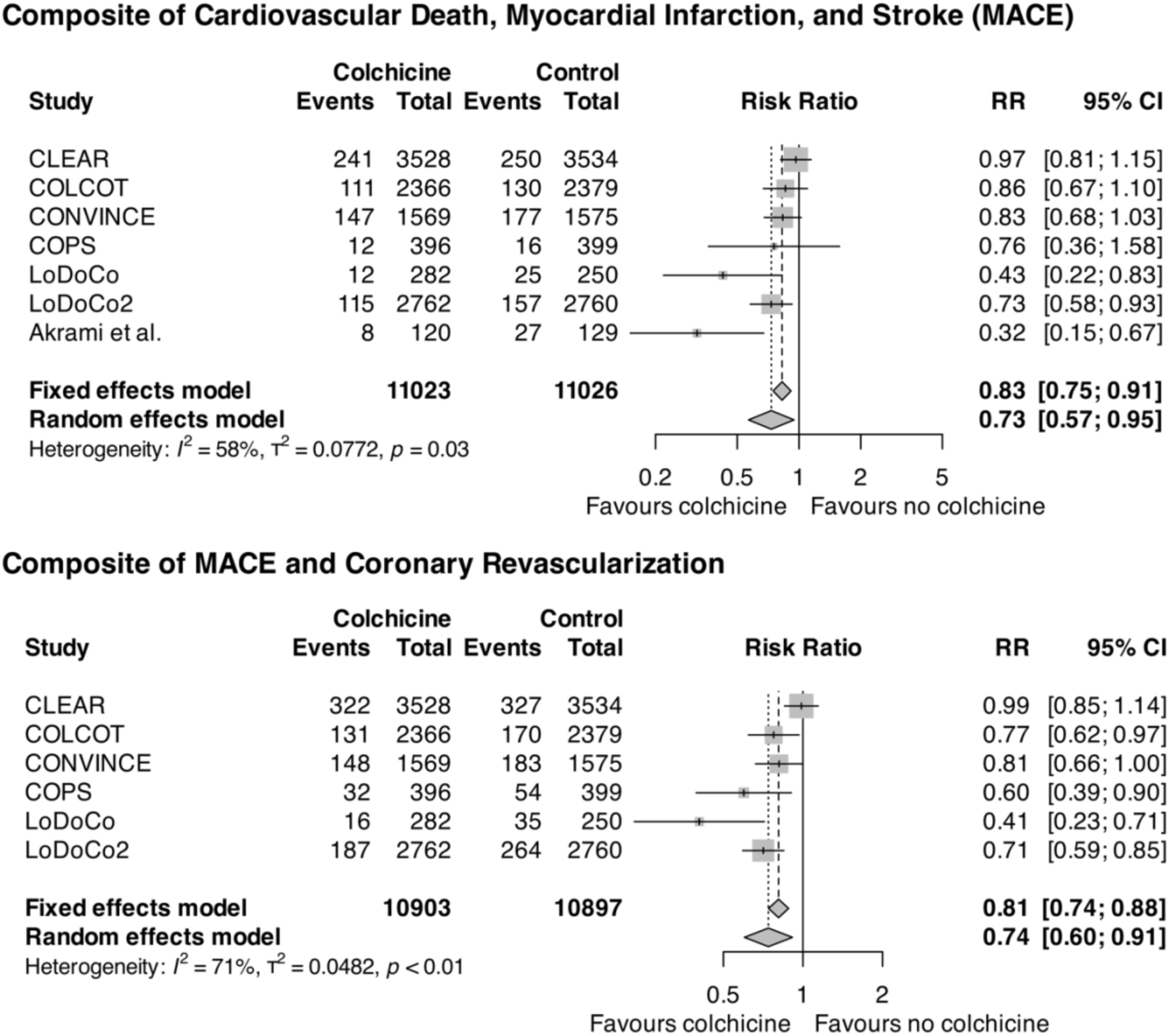
Cardiovascular composite outcomes (MACE with and without coronary revascularization) in patients treated with colchicine in addition to standard of care versus standard of care alone. RR = Risk Ratio, CI = Confidence Interval.

At Galbraith plot analysis, three trials were identified as the main outliers, with CLEAR[22] showing a neutral effect of colchicine, while LoDoCo[30] and Akrami et al.[28] showed a RR of 0.43 and 0.32, respectively. Funnel plot inspection did not show clear publication bias, suggesting only mild asymmetry in small studies (**Supplementary Figure S3**).

Meta regression showed that active smoking significantly affected the effect of colchicine (β = 1.10, 95% CI 0.21 - 1.99, p = 0.016), with a greater treatment effect observed in populations with lower smoking rates. Other covariates did not affect the treatment effect.

Subgroup analysis of trials enrolling ACS vs CCS patients showed a neutral effect of colchicine (RR 0.73, 95% CI 0.47 – 1.14) in the ACS group, but there were no significant differences with the CCS group (p = 0.60).

### Secondary outcomes

Colchicine reduced the relative risk of the composite of MACE and coronary revascularization by 26% (RR 0.74, 95% CI 0.60 - 0.91, I^2^ = 71%). Heterogeneity was mainly driven by CLEAR[22] showing neutral results (RR 0.99, 95% CI 0.85 - 1.14), while COLCOT and LoDoCo showed a RR 0.77 and 0.71, respectively.[17,30] Among individual components (**Figure 1**, **Figure 3**), colchicine significantly reduced the risk of myocardial infarction (RR 0.83, 95% CI 0.72 - 0.96, I² = 0%) and coronary revascularization (RR 0.79, 95% CI 0.65 - 0.94, I² = 30%). A trend towards stroke reduction was observed but did not reach statistical significance (RR 0.73, 95% CI 0.50 - 1.06). Cardiovascular mortality was similar between groups (RR 0.97, 95% CI 0.79 - 1.19; **Figure 4**). Results for individual endpoints were relatively homogeneous across all trials (I² < 50%). Funnel plot inspection did not show clear publication bias, suggesting only mild asymmetry in small studies (**Supplementary Figure S3**). Meta-regression analyses did not identify any significant effect modifiers for these outcomes. The estimated number needed to treat to avoid one occurrence of myocardial infarction or unplanned coronary revascularization were 168 and 92, respectively. Subgroup analysis of trials enrolling ACS vs CCS patients showed a neutral effect of colchicine on all secondary outcomes in the ACS group, but there were no significant differences with the CCS group.

**Figure 3.**
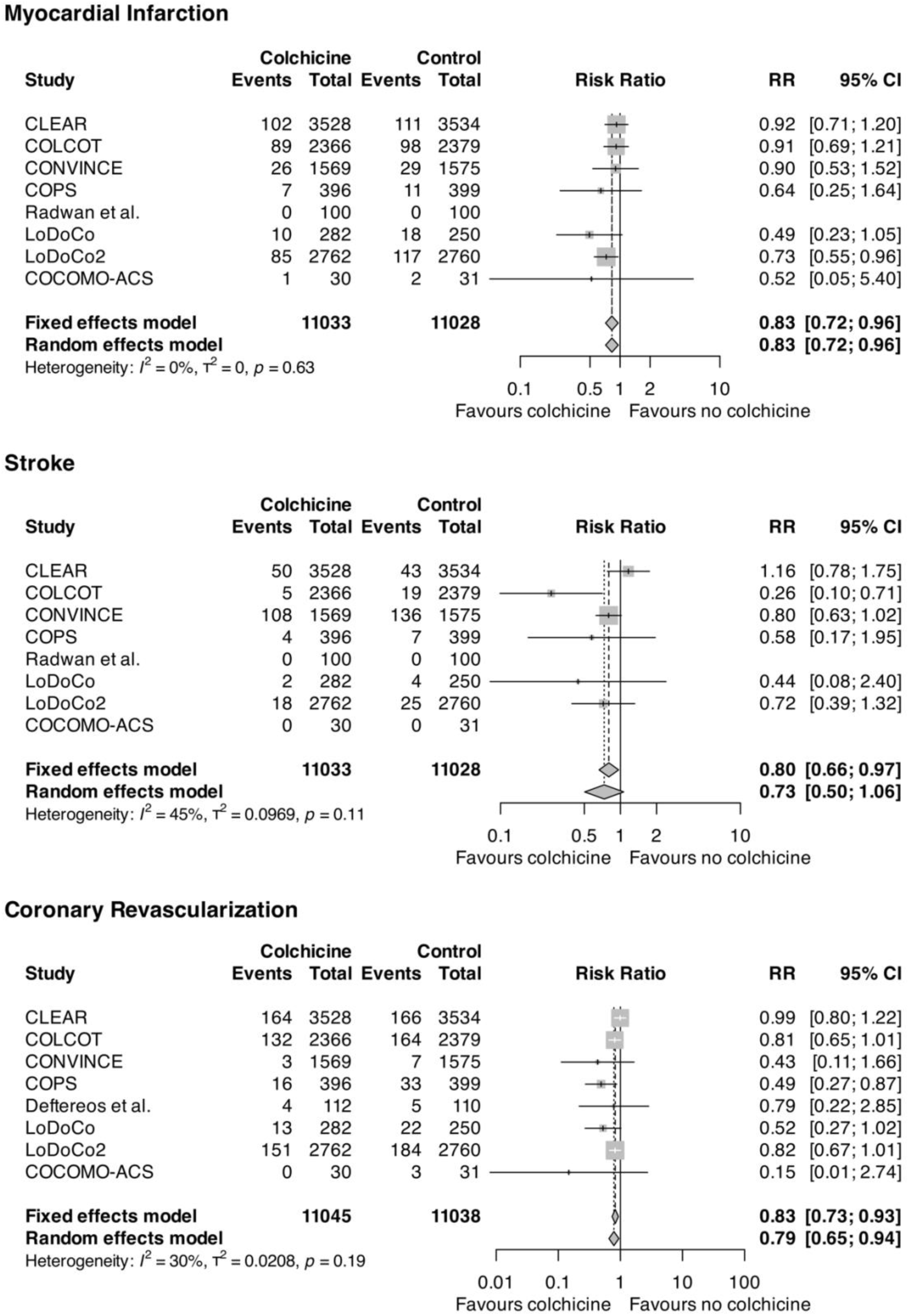
Myocardial Infarction, Stroke and Coronary Revascularization in patients treated with colchicine in addition to standard of care versus standard of care alone. RR = Risk Ratio, CI = Confidence Interval.

**Figure 4.**
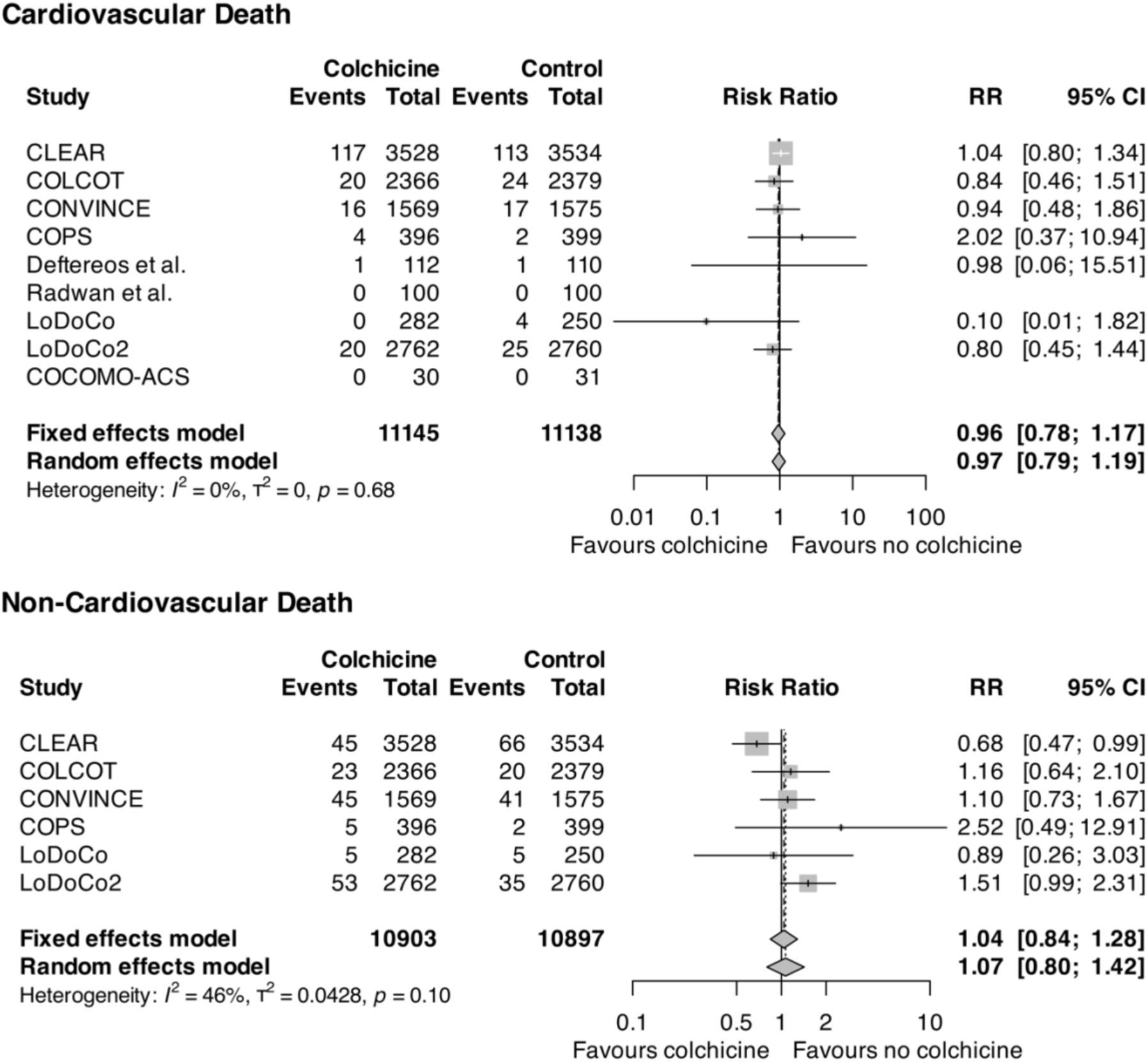
Death in patients treated with colchicine in addition to standard of care versus standard of care alone. RR = Risk Ratio, CI = Confidence Interval.

### Safety Outcomes

Colchicine therapy did not increase the risk of non-cardiovascular mortality (RR 1.07, 95% CI 0.80 - 1.42, I² = 46%; **Figure 4**). The risk of serious infections was similar between groups (RR 0.99, 95% CI 0.79 - 1.25, I² = 36 %). Serious gastrointestinal adverse events showed a slight numerical increase with colchicine, although not reaching statistical significance (RR 1.13, 95% CI 0.92 - 1.39, I² = 0%). For pneumonia requiring hospitalization, while the pooled estimate showed no significant difference (RR 1.33, 95% CI 0.60 - 2.95), there was substantial heterogeneity across trials (I² = 64%), with COLCOT[17] showing an increased risk (RR 2.35, 95% CI 1.08 - 5.11) not confirmed in other studies. Importantly, cancer incidence was similar between groups (RR 0.97, 95% CI 0.82 - 1.15, I² = 0%). Forest plots for these safety outcomes are shown in **Supplementary Figure S4**.

## Discussion

This study-level meta-analysis of ten randomized clinical trials enrolling 22532 patients with ASCVD provides evidence that long-term low-dose colchicine is effective in reducing the risk of MACE, myocardial infarction and unplanned coronary revascularization in patients with ASCVD.

Previous meta-analyses have shown a protective effect of colchicine when compared to placebo or standard of treatment in regard to MACE, stroke, myocardial infarction and coronary revascularization.[33,34]

The recent publication of CLEAR,[22] the largest trial on long-term colchicine therapy in patients with ASCVD, challenged these findings showing an overall neutral effect of colchicine in acute coronary syndromes on the primary outcome of MACE.

To the best of our knowledge, this meta-analysis is the first to pool the effect of the CLEAR trial with other landmark trials on long-term low-dose colchicine in ASCVD. Regarding the primary endpoint (MACE), CLEAR[22] was the one with the highest relative weight among the included studies (32.0%, followed by CONVINCE[21] and LoDoCo2[18] with 22.6% and 20.1%, respectively). Nonetheless, this meta-analysis demonstrates a significant 27% relative risk reduction in MACE with colchicine therapy, with an estimated number needed to treat of 52 patients to prevent one event, which is consistent with previous reports while considering a substantially larger population.

However, it should be noted that including CLEAR lead to a meaningful increase in the study heterogeneity, which requires careful consideration.

While both trials on long-term low-dose colchicine in CCS (LoDoCo and LoDoCo2) [18,30] were positive for a reduction of MACE in patients in the experimental group, trials enrolling patients with ACS have mostly been neutral, with the notable exception of a relatively smaller trial by Akrami and colleagues.[28] While subgroup analysis showed a neutral pooled effect of colchicine in ACS patients (RR 0.73, 95% CI 0.47 – 1.14), there was no statistically significant interaction between clinical presentation subgroups (p = 0.60).

These findings suggest that the presence of benefit of colchicine in ACS patients cannot be excluded with the available evidence, and the treatment effect should be considered consistent across clinical presentations until definitively proven otherwise by future research.

The identification of smoking as a significant effect modifier (β = 1.10, p = 0.016) requires careful consideration. When considering the trials used for meta regression, the heterogeneity in the proportion of patients with active smoking appears to be quite significant, ranging from 4.5% and 11.8% (LoDoCo[30] and LoDoCo2[18] trials, respectively) to 40.8% (CLEAR)[22].

While providing a definite mechanistic explanation on the complex interplay between active smoking and colchicine is beyond the scope of this systematic review, the finding appears biologically plausible. Colchicine works by inhibiting microtubule polymerization, which reduces the activity of neutrophils and other inflammatory cells.[1,35] On the other hand, active smoking is known to promote chronic systemic and vascular inflammation through oxidative stress. Notably, active smoking may lead to cellular contraction and degeneration mediated by specifically interfering with microtubule function, possibly undermining a critical pharmacodynamic target in long term colchicine therapy.[36]

Our meta-analysis confirms the reduction in the risk of myocardial infarction and unplanned coronary revascularization in patients in the colchicine group, by 17% and 21% respectively. On the other hand, the inclusion of the CLEAR trial[22] in the pooled analysis for individual outcomes challenges the previously reported reduction in the risk of stroke. While less events were reported in the colchicine group [187 (risk: 1.7%) versus 234 (risk: 2.1%)], this finding did not reach statistical significance (RR 95% CI 0.50 – 1.06). Notably, the omission of CLEAR at leave-one-out analysis shows a significant reduction of 28% for the risk of stroke (95% CI 0.58 – 0.89). Interestingly, the outcome definition of Stroke reported by the CLEAR trial[22] uniquely includes subarachnoid haemorrhage and does not require imaging (CT or MRI) confirmation, potentially incorporating non-ischemic events that could dilute the treatment effect.

Regarding the effects on mortality and the safety outcomes, the present results align with previously published meta-analytic reports.[37] Nonetheless, the included trials were not powered to assess differences in these respect and the known side-effect spectrum of colchicine and concerns regarding the long-term immunomodulatory effects warrants careful evaluation of pooled data. While our meta-analysis provides reassuring data regarding the safety of low-dose colchicine treatment for all the safety outcomes, trials were not designed with appropriate power or follow-up time to thoroughly assess differences in mortality. Interestingly, all trials reported a higher rate of non-cardiovascular death compared to cardiovascular death, except for CLEAR,[22] despite the high cardiovascular risk of the enrolled populations. While this finding has been previously reported in published literature as well as recent meta-analyses,[33,34,38] CLEAR’s divergent finding warrants careful interpretation, particularly given the size and the long follow-up period. Notably, the trial’s definition of cardiovascular death included deaths of unknown cause, and was conducted during the COVID-19 pandemic without specific reporting of respiratory-related hospitalizations, with potential misclassification of events.

The Food and Drug Administration (FDA) has approved the use of low dose colchicine for secondary prevention in ischemic heart disease,[39] while the recent 2024 European Society of Cardiology (ESC) guidelines on chronic coronary syndromes[40] have upgraded its recommendation to class IIA from previous 2021 ESC guidelines on cardiovascular prevention.[41] This systematic review provides evidence that combined results of available clinical trials suggest that colchicine effectively reduces MACE in patients with acute and especially chronic coronary syndromes, even after the neutral results of the CLEAR trial.

### Limitations

This meta-analysis presents several limitations. Trial differences in population and outcome definition introduced heterogeneity in the pooled estimates, and reported data did not allow for time-to-event analysis, which was planned as a secondary analysis in the prospective analysis plan. Only six of the included trials provided clear reporting on all the pre-specified outcomes and predictors. Moreover, this analysis used study-level data, that carries a risk of losing important information without access to individual patient data.

## Conclusions

The addition of colchicine reduces the risk of MACE, myocardial infarction and coronary revascularization in patients with ASCVD treated with standard of therapy, with a good safety profile. Our findings support the use of long-term low-dose colchicine for secondary prevention of MACE in clinical practice.

## Patient and public involvement

Patients and/or the public were not involved in the design, or conduct, or reporting, or dissemination plans of this research.

## Data availability statement

Data are available on reasonable request.

## Data Availability

All data produced in the present study are available upon reasonable request to the authors

