## Supplemental material for "Colchicine for Prevention of Major Adverse Cardiovascular Events: A systematic review and meta-analysis of randomized clinical trials"

### Supplementary Table S1: PRISMA checklist

| Section and Topic | Item # | Checklist item | Location where item is reported |
|---|---|---|---|
| **TITLE** | | | |
| Title | 1 | Identify the report as a systematic review. | Title, 73 |
| **ABSTRACT** | | | |
| Abstract | 2 | See the PRISMA 2020 for Abstracts checklist. | 32-52 |
| **INTRODUCTION** | | | |
| Rationale | 3 | Describe the rationale for the review in the context of existing knowledge. | 68-74 |
| Objectives | 4 | Provide an explicit statement of the objective(s) or question(s) the review addresses. | 68-74 |
| **METHODS** | | | |
| Eligibility criteria | 5 | Specify the inclusion and exclusion criteria for the review and how studies were grouped for the syntheses. | 91-95 |
| Information sources | 6 | Specify all databases, registers, websites, organisations, reference lists and other sources searched or consulted to identify studies. Specify the date when each source was last searched or consulted. | 80-89 |
| Search strategy | 7 | Present the full search strategies for all databases, registers and websites, including any filters and limits used. | 80-89 |
| Selection process | 8 | Specify the methods used to decide whether a study met the inclusion criteria of the review, including how many reviewers screened each record and each report retrieved, whether they worked independently, and if applicable, details of automation tools used in the process. | 96-99 |
| Data collection process | 9 | Specify the methods used to collect data from reports, including how many reviewers collected data from each report, whether they worked independently, any processes for obtaining or confirming data from study investigators, and if applicable, details of automation tools used in the process. | 96-99 |
| Data items | 10a | List and define all outcomes for which data were sought. Specify whether all | 107-113 |

| Section and Topic | Item # | Checklist item | Location where item is reported |
|---|---|---|---|
| | | results that were compatible with each outcome domain in each study were sought (e.g. for all measures, time points, analyses), and if not, the methods used to decide which results to collect. | |
| | 10b | List and define all other variables for which data were sought (e.g. participant and intervention characteristics, funding sources). Describe any assumptions made about any missing or unclear information. | 99-100 |
| Study risk of bias assessment | 11 | Specify the methods used to assess risk of bias in the included studies, including details of the tool(s) used, how many reviewers assessed each study and whether they worked independently, and if applicable, details of automation tools used in the process. | 100-105 |
| Effect measures | 12 | Specify for each outcome the effect measure(s) (e.g. risk ratio, mean difference) used in the synthesis or presentation of results. | 115-116 |
| Synthesis methods | 13a | Describe the processes used to decide which studies were eligible for each synthesis (e.g. tabulating the study intervention characteristics and comparing against the planned groups for each synthesis (item #5)). | Supplementary Table S1 |
| | 13b | Describe any methods required to prepare the data for presentation or synthesis, such as handling of missing summary statistics, or data conversions. | Not applicable |
| | 13c | Describe any methods used to tabulate or visually display results of individual studies and syntheses. | Figures 2-4 |
| | 13d | Describe any methods used to synthesize results and provide a rationale for the choice(s). If meta-analysis was performed, describe the model(s), method(s) to identify the presence and extent of statistical heterogeneity, and software package(s) used. | 115-127 |
| | 13e | Describe any methods used to explore possible causes of heterogeneity among study results (e.g. subgroup analysis, meta-regression). | 115-118, 119-124 |
| | 13f | Describe any sensitivity analyses conducted to assess robustness of the synthesized results. | 116-117, 123-124 |
| Reporting bias assessment | 14 | Describe any methods used to assess risk of bias due to missing results in a synthesis (arising from reporting biases). | 124-125 |

| Section and Topic | Item # | Checklist item | Location where item is reported |
|---|---|---|---|
| Certainty assessment | 15 | Describe any methods used to assess certainty (or confidence) in the body of evidence for an outcome. | 124-125 |
| **RESULTS** | | | |
| Study selection | 16a | Describe the results of the search and selection process, from the number of records identified in the search to the number of studies included in the review, ideally using a flow diagram. | Supplementary Figure S1 |
| | 16b | Cite studies that might appear to meet the inclusion criteria, but which were excluded, and explain why they were excluded. | Supplementary Figure S1 |
| Study characteristics | 17 | Cite each included study and present its characteristics. | Table 1, 139-155 |
| Risk of bias in studies | 18 | Present assessments of risk of bias for each included study. | Supplementary Figure S2 |
| Results of individual studies | 19 | For all outcomes, present, for each study: (a) summary statistics for each group (where appropriate) and (b) an effect estimate and its precision (e.g. confidence/credible interval), ideally using structured tables or plots. | Figures 2-4 |
| Results of syntheses | 20a | For each synthesis, briefly summarise the characteristics and risk of bias among contributing studies. | 140-187 |
| | 20b | Present results of all statistical syntheses conducted. If meta-analysis was done, present for each the summary estimate and its precision (e.g. confidence/credible interval) and measures of statistical heterogeneity. If comparing groups, describe the direction of the effect. | 140-187 |
| | 20c | Present results of all investigations of possible causes of heterogeneity among study results. | 140-187 |
| | 20d | Present results of all sensitivity analyses conducted to assess the robustness of the synthesized results. | 140-187 |
| Reporting biases | 21 | Present assessments of risk of bias due to missing results (arising from reporting biases) for each synthesis assessed. | Not applicable |
| Certainty of | 22 | Present assessments of certainty (or confidence) in the body of evidence for each | 140-197 |

| Section and Topic | Item # | Checklist item | Location where item is reported |
|---|---|---|---|
| evidence | | outcome assessed. | |
| **DISCUSSION** | | | |
| Discussion | 23a | Provide a general interpretation of the results in the context of other evidence. | 200-271 |
| | 23b | Discuss any limitations of the evidence included in the review. | 273-278 |
| | 23c | Discuss any limitations of the review processes used. | 273-278 |
| | 23d | Discuss implications of the results for practice, policy, and future research. | 280-283 |
| **OTHER INFORMATION** | | | |
| Registration and protocol | 24a | Provide registration information for the review, including register name and registration number, or state that the review was not registered. | 77-78 |
| | 24b | Indicate where the review protocol can be accessed, or state that a protocol was not prepared. | 77-78 |
| | 24c | Describe and explain any amendments to information provided at registration or in the protocol. | 77-78 |
| Support | 25 | Describe sources of financial or non-financial support for the review, and the role of the funders or sponsors in the review. | Not applicable |
| Competing interests | 26 | Declare any competing interests of review authors. | 288 |
| Availability of data, code and other materials | 27 | Report which of the following are publicly available and where they can be found: template data collection forms; data extracted from included studies; data used for all analyses; analytic code; any other materials used in the review. | 288 |

425

**Supplementary Table S2. Complete inclusion and exclusion criteria of the systematic review and meta-analysis**

| **Inclusion criteria** | **Exclusion criteria** |
|---|---|
| Randomized clinical trials | Absence of reported outcomes for cardiovascular death or myocardial infarction |
| Adult participants with established atherosclerotic disease, including coronary artery disease, peripheral artery disease, or cerebrovascular disease; | Conference abstracts |
| Intervention consisting of long-term colchicine treatment (>3 months) compared to standard treatment with or without placebo. | |

**Supplementary Table S3: Medical therapy at randomization. DAPT = Dual antiplatelet therapy; ACEi/ARB = angiotensin-converting enzyme inhibitors/angiotensin receptor blocker; NA = not available. Data on medical therapy at randomization not available for Deftereos et al, Akrami et al, Radwan et al.**

| | Antiplatelet agents | | | | Statins (%) | Beta-blockers (%) | ACEi/ARB (%) | SGLT2 inhibitors |
|---|---|---|---|---|---|---|---|---|
| | Any (%) | Aspirin (%) | P2Y12 inhibitors (%) | DAPT (%) | | | | |
| CONVINCE | 97.5 | NA | NA | NA | 93.8 | NA | NA | NA |
| CLEAR | NA | 96.8 | 85.7 | NA | 96.6 | NA | 78.1 | 3 |
| COCOMO-ACS | 100 | NA | NA | NA | 95.3 | 57.8 | 70.3 | NA |
| COPS | NA | 98.6 | NA | 97.1 | 98.9 | 82.6 | 86.9 | NA |
| LoDoCo2 | 67 | NA | NA | 23.2 | 94 | 62.1 | 71.7 | NA |
| COLCOT | NA | 98.8 | 97.9 | NA | 99 | 88.9 | NA | NA |
| LoDoCo | 93.4 | NA | NA | 11.7 | 95.1 | 66.5 | 29.1 | NA |

**Supplementary Table S4: Coronary syndrome trials – qualifying events (acute versus chronic coronary syndromes). ACS = Acute coronary syndrome; CCS = chronic coronary syndrome.**

| | ACS (n) | CCS (n) |
|---|---|---|
| CLEAR | 7062 | 0 |
| COCOMO-ACS | 64 | 0 |
| Radwan et al. | 200 | 0 |
| COPS | 773 | 0 |
| Akrami et al. | 249 | 0 |
| LoDoCo2 | 0 | 5522 |
| COLCOT | 4745 | 0 |
| LoDoCo | 0 | 532 |
| Deftereos et al. | 61 | 135 |

**Supplementary Table S5: Coronary syndrome trials stratifying by Myocardial Infarction and Unstable Angina definition. MI = myocardial infarction; UA= unstable angina.**

| | MI<br>n (%) | UA<br>n (%) |
|---|---|---|
| CLEAR | 7062 (100) | 0 |
| COCOMO-ACS | 64 (100) | 0 |
| Radwan et al. | 200 (100) | 0 |
| COPS | 747 (96.7) | 26 (3.3) |
| Akrami et al. | 163 (65.5) | 86 (34.5) |
| COLCOT | 4745 (100) | 0 |

**Supplementary Table S6: Trials enrolling ACS patients divided by ACS type. ACS = acute coronary syndrome; STEMI = ST-elevation myocardial infarction; NSTEMI = non-ST-elevation myocardial infarction; UA = unstable angina.**

| | STEMI<br><br>n (%) | NSTEMI<br><br>n (%) | UA<br><br>n (%) |
|---|---|---|---|
| CLEAR | 349 (4.9) | 6713 (95.1) | 0 |
| COCOMO-ACS | 0 | 64 (100) | 0 |
| Radwan et al. | 200 (100) | 0 | 0 |
| COPS | 390 (49.1) | 357 (44.9) | 26 (3.3) |
| Akrami et al. | 128 (51.4) | 35 (14.1) | 86 (34.5) |

**Supplementary Figure S1. PRISMA flow diagram**

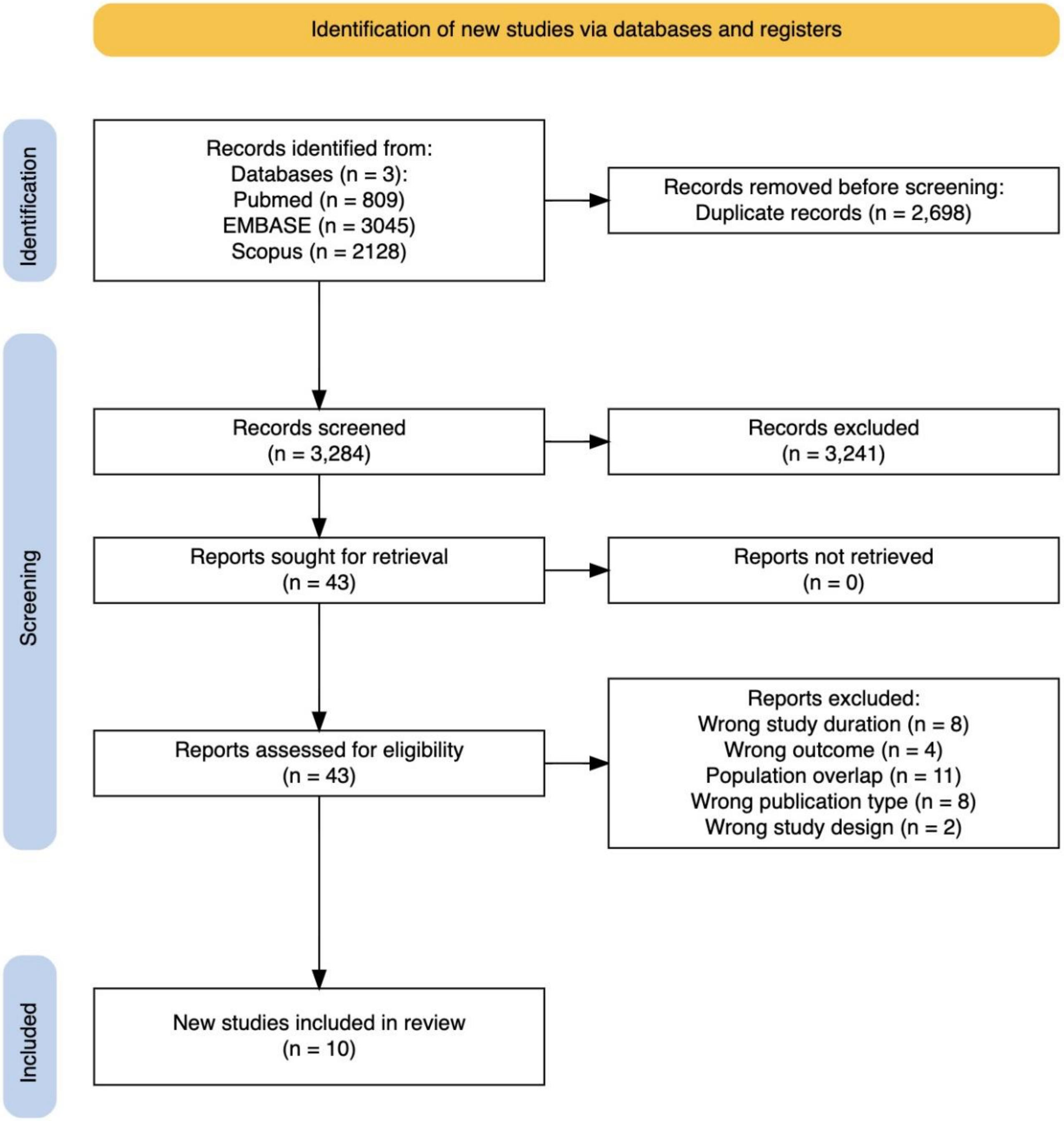

**Supplementary Figure S2. Risk of bias for the included trials according to the revised risk-of-bias tool for randomized trial (Rob2 version 2.0)**

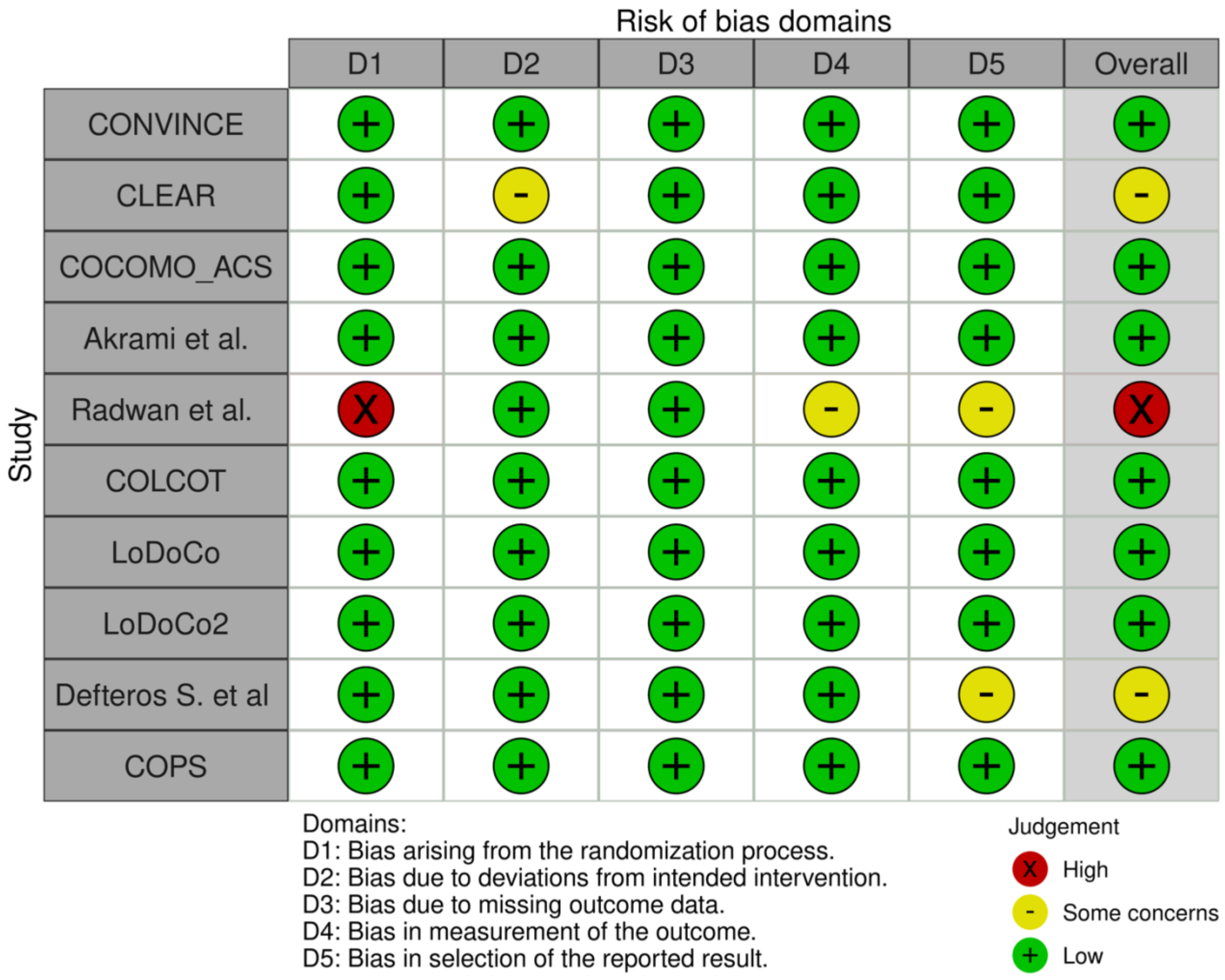

**Supplementary Figure S3. Funnel plots for the reported efficacy outcomes.**

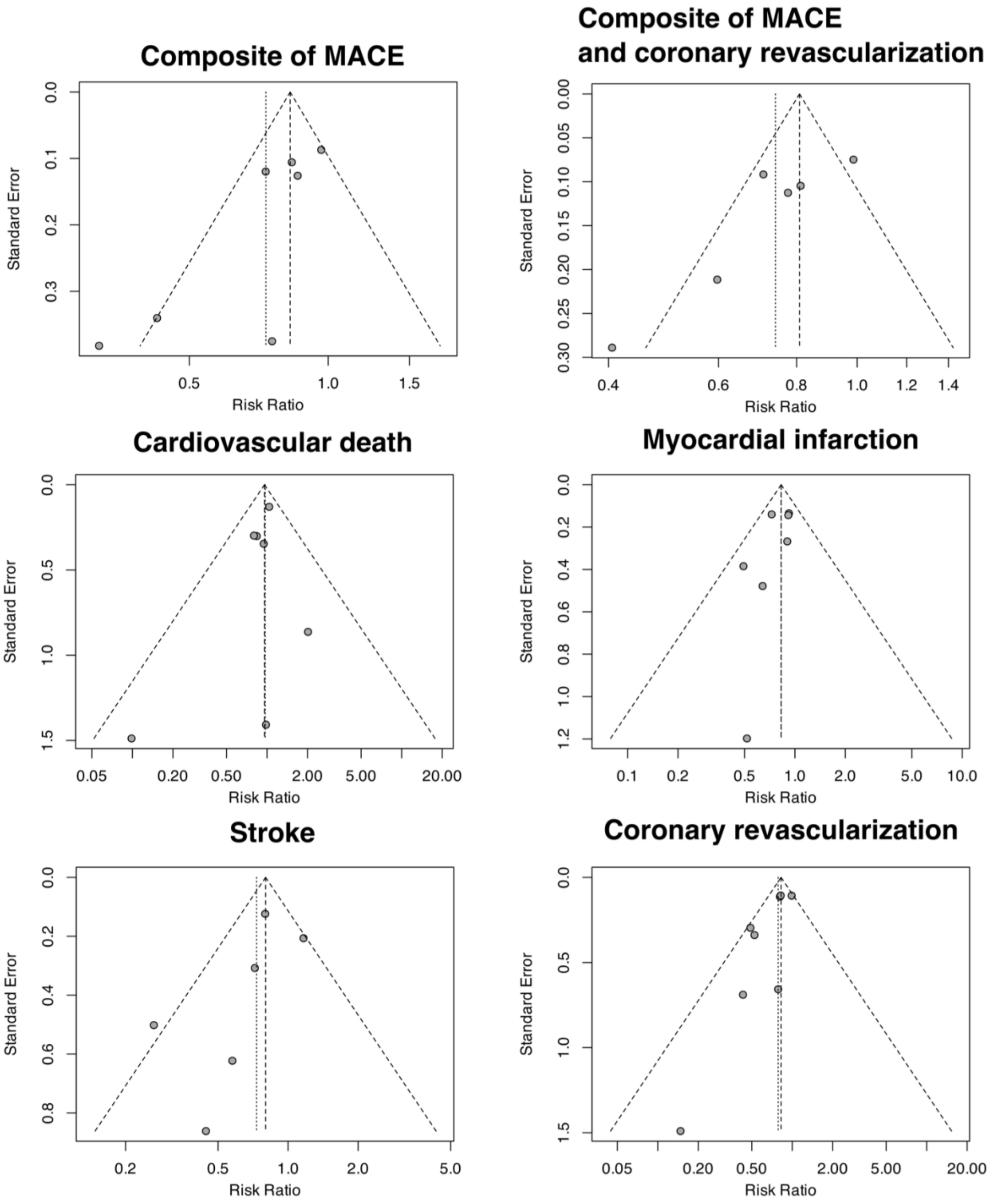

**Supplementary Figure S4. Safety outcomes in patients treated with colchicine in addition to standard of care versus standard of care alone. RR = Risk Ratio, CI = Confidence Interval.**

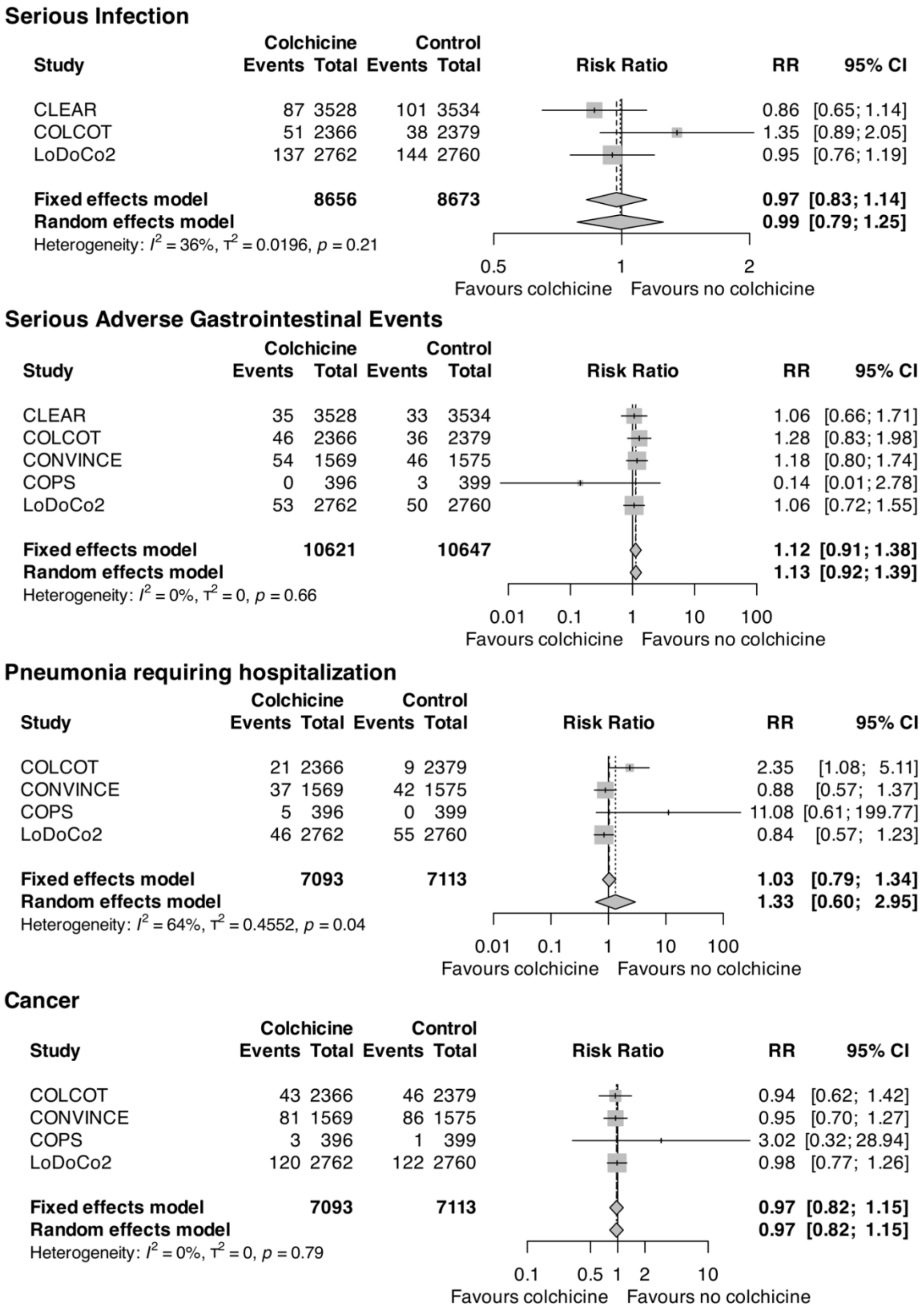
